# Differences Between Brachial And Aortic Blood Pressure In Adolescence and their implications for diagnosis of hypertension

**DOI:** 10.1101/2023.10.10.23296853

**Authors:** Alun D Hughes, George Davey Smith, Laura D Howe, Deborah Lawlor, Siana Jones, Chloe M Park, Nish Chaturvedi

## Abstract

**Objectives:** Blood pressure is the leading global cause of mortality, and its prevalence is increasing in children and adolescents. Aortic blood pressure (BP) is lower than brachial BP in adults. We aimed to assess the extent of this difference and its impact on the diagnosis of hypertension among adolescents.

**Methods:** We used data from 3850 participants from a UK cohort of births in the early 1990s in the Southwest of England, who attended their ∼17 year follow-up and had valid measures of brachial and aortic BP at that clinic (mean(SD) age 17.8(0.4)y, 66% female). Data are presented as mean differences [95% prediction intervals] for both sexes.

**Results:** Aortic systolic BP was lower than brachial systolic BP (male, -22.3[-31.2, - 13.3]mmHg; female, -17.8[-25.5, -10.0]mmHg). Differences between aortic and brachial diastolic BP were minimal. Based on brachial BP measurements, 101 males (6%) and 22 females (1%) were classified as hypertensive. In contrast, only nine males (<1%) and 14 females (<1%) met the criteria for hypertension based on aortic BP, and the predictive value of brachial BP for aortic hypertension was poor (positive predictive value = 13.8%). Participants with aortic hypertension had a higher left ventricular mass index than those with brachial hypertension.

**Conclusions:** Brachial BP substantially overestimates aortic BP in adolescents due to marked aortic-to-brachial pulse pressure amplification. The use of brachial BP measurement may result in an overdiagnosis of hypertension during screening in adolescence.

## Introduction

High blood pressure (BP) is the leading global cause of mortality.[1] Elevated BP is frequently evident in youth, with recent studies reporting that around 2 to 13% of young people may be hypertensive, depending on geographical region and definition used to classify hypertension.[2,3] Disturbingly, there is evidence that the prevalence of elevated BP in children is increasing,[4] probably as a result of the global epidemic of obesity and physical inactivity.[5] Elevated BP in young people is important as there is evidence that BP tracks into adulthood,[6] and has long term implications for cardiovascular health and mortality.[7,8] Conversely, a diagnosis of hypertension can have adverse psychological consequences[9] and pharmacological treatment for hypertension has adverse effects. This is particularly relevant because the benefits and harms of antihypertensive medication have not been studied extensively in young people.[2,10]

It is well recognised that systolic BP can differ substantially depending on the site of measurement. Typically brachial systolic BP is higher than aortic systolic BP because of pulse pressure amplification due to wave reflection.[11] In adults pulse pressure amplification is highly variable,[12] tends to decrease with age[13] and can account for isolated systolic hypertension (ISH).[14] ISH in adults between 18 and 39 years is more common than systolic plus diastolic hypertension,[15] although its clinical significance is debated in this age-group.[16,17] Further, there is evidence that aortic as opposed to brachial BP is a more relevant prognostic indicator - being more strongly associated with cardiovascular events in adults,[18,19] and with more target organ damage in both adults[20] and adolescents.[21]

At present, there is limited evidence on the extent of the difference between aortic and brachial BP in adolescents, the prevalence of isolated systolic hypertension in people below the age of 18, or its impact on the diagnosis of hypertension in youth, an issue highlighted by recent international guidelines.[3] We therefore, we aimed to determine the difference in 1) brachial and aortic BP in a large sample of adolescents drawn from an English birth cohort, the Avon Longitudinal Study of Parents and Children (ALSPAC); and 2) the proportion of people with hypertension or ISH in this sample based on the use of brachial or aortic BP. A further aim was to determine whether aortic and brachial blood pressure, hypertension, and ISBP differed in relation to target organ damage to the heart (i.e., left ventricular mass index (LVMI)) or vasculature (i.e., carotid intima media thickness (cIMT) and carotid-femoral pulse wave velocity (cfPWV)), and to investigate possible factors that contributed to aortic to brachial pulse pressure amplification in adolescents.

## Methods

### Study design and participants

Pregnant women resident in the former county of Avon, Southwest England, with expected dates of delivery from April 1, 1991, to December 31, 1992 were invited to participate in The Avon Longitudinal Study of Parents and Children (ALSPAC). The initial number of pregnancies enrolled was 14,541 (for these, at least one questionnaire was returned or a “Children in Focus” clinic was attended by 19/07/99). Of these initial pregnancies, there were 14,062 live births and 13,988 children who were alive at 1 year of age and have been followed since then.[22,23] The study website (http://www.bristol.ac.uk/alspac/researchers/our-data/) contains details of all available data. The present analysis was based on 5,081 participants aged ∼17 years who attended the ALSPAC F17 clinic between 2009 and 2011 as part of an ongoing follow-up. Ethical approval was obtained from the ALSPAC Law and Ethics Committee and Local Research Ethics Committee, and all participants provided written informed consent.

Individuals with diabetes mellitus (n=21), familial hypercholesterolemia (n=8), known heart disease (n=3), pregnancy (n=15), or those who did not participate in the BP measurement session for any reason (n = 741) were excluded. A further 11 people refused tonometry measurements, and in 392 people tonometry measurements failed quality control procedures, yielding a total of 3850 evaluable recordings (Supplementary Figure S1).

### Clinic measurements

Patient age and sex were recorded at the clinic. Demographic and lifestyle data were obtained using a questionnaire. Socioeconomic position was assessed based on the father’s occupation (using the 1991 UK Office of Population Censuses and Surveys classification) and mother’s education ((less than O-level, O-level, or more than O-level but no degree, and degree or above, where O-levels were the standard school-leaving qualifications taken around age 16 years until recently in the UK). Alcohol consumption was assessed as the number of drinks containing alcohol consumed on a typical day, and smoking was categorized as never, ever but not currently, or currently. Weight and height were measured while the participants wore light clothing and no shoes. Height was estimated to the nearest 0.1 cm with a Harpenden Stadiometer. Body weight was measured to the nearest 0.1 kg using a Tanita TBF 305 scale. Body composition was assessed using a Lunar Prodigy Dual-energy X-ray absorptiometry scanner (GE Medical Systems, Madison, Wisconsin, USA). Habitual physical activity was assessed using a hip-worn uniaxial ActiGraph device between the age 14-17 (AM7164 2.2; ActiGraph LLC, Fort Walton Beach, FL, USA) and average daily minutes of total physical activity at light, moderate or vigorous intensity was calculated based on cut-points of 200–3,599, 3,600–6,199 and ≥6,200 cpm, respectively. Carotid femoral pulse wave velocity (cfPWV) was measured using a Vicorder device (SMT Medical Technology GmbH, Bristol, UK) as previously described.[24] Carotid intima media thickness (cIMT) was measured in left and right common carotid arteries by ultrasound using a linear 12-MHz transducer (Vivid7, GE Medical, Chicago, Illinois) and averaged.[25] Left ventricular mass was measured in approximately 1 in 2 participants selected in a quasi-random fashion using an ultrasound device (HDI 5000, Philips Healthcare, North Andover, MA, USA) equipped with a P4-2 Phased Array ultrasound transducer according to the American Society of Echocardiography guidelines, as previously described and indexed to height^1.7^.[26] Blood samples were collected following an overnight fast for those assessed in the morning or a minimum of 6 hours fasting for those assessed in the afternoon. Samples were centrifuged immediately, separated, and frozen at -80°C before analysis. Lipid profiles (total cholesterol, high-density lipoprotein (HDL) cholesterol, triglycerides), glucose, and insulin were measured as described previously.[27] Insulin sensitivity (HOMA-S) was estimated using the Homeostasis Assessment Model (Version 2.2.3).[28]

The participants’ sitting BP and heart rate were measured at least three times with at least a minute interval using an Omron 705IT device according to contemporary guidelines in the dominant arm using an appropriate cuff size.[29] The average of the final two readings was used. The BP waveform was measured using radial tonometry (SphygmoCor, AtCor Medical), and aortic pressure was estimated using a generalized transfer function (GTF), which has been validated in adults and children.[12,30] The late systolic shoulder (SBP2) was also used as an alternative estimate of aortic systolic BP, which does not rely on a GTF.[31] Amplification was calculated as brachial pulse pressure/aortic pulse pressure. All measurements were made by trained investigators and ongoing quality control was conducted throughout the study; reproducibility was excellent, as has been reported previously.[32]

In accordance with ESH guidelines,[33] hypertension was defined as brachial BP ≥140/90 mmHg. Isolated systolic hypertension (ISH) was defined as systolic BP ≥140 mmHg and diastolic BP <90 mmHg.[33] For the classification of hypertension based on aortic BP, we used the definition of aortic BP ≥130/90 mmHg as hypertensive,[34,35] and aortic systolic BP ≥130 and diastolic BP <90 mmHg as indicative of aortic ISH (aISH). A subsidiary analysis using the recent American Academy of Pediatrics Clinical Practice Guideline recommendations[2] (Stage I hypertension: ≥130/80mmHg) was also performed.

### Statistical analysis

All analyses were performed in Stata/MP 17.0 (StataCorp). Descriptive statistics for continuous variables are presented as mean (SD), and N(%) for categorical variables. Comparisons between the included sample and those eligible but not included were made using Student’s t-tests or Chi^2^ tests as appropriate. Analyses were stratified by sex, based on previous evidence of sex differences in the aortic BP waveform and amplification.[36] For the main analysis of difference in brachial and aortic BP we used unadjusted linear regression or 2-dimension fractional polynomial if there was evidence of non-linearity. If there was evidence of heteroscedasticity, standard errors for the linear model were estimated using the bootstrap estimator. Linear regression results were summarized as beta coefficients with 95% confidence intervals (95% CI). When nonlinearity and heteroskedasticity were present, we used a combination of fractional polynomials and quantile regression to produce median, 5%, and 95% quantile boundaries for the nonlinear relationship [95% QI]. For differences in the proportions of different definitions of HR using aortic and brachial BP, we used unadjusted logistic regression. We estimated the sensitivity, specificity, and positive and negative prediction metrics for hypertension and ISH by using aortic and brachial BP. Associations between aortic or brachial hypertension in the absence of aortic hypertension and measures of target organ damage were adjusted for potential confounders (age, BMI, fat and lean mass, habitual physical activity, maternal education, and socioeconomic status) chosen on the basis of background knowledge. To investigate potential predictors of AMPLIFICATION, we used a linear model with inclusion of clinical predictors (see Table 1), and model selection was performed using an elastic net with 10-folds. We report the % variation explained in the outcome for all selected variables.

**Table 1.**
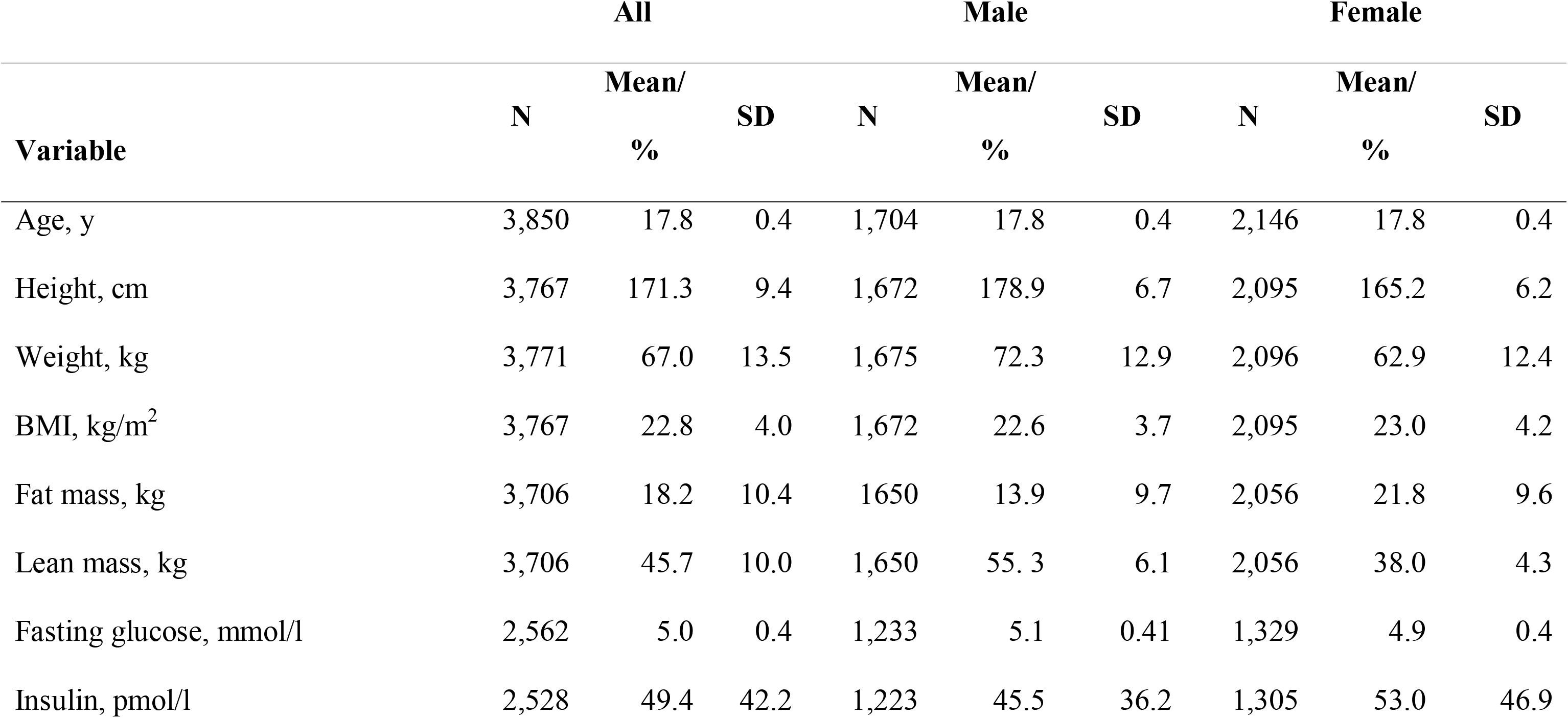
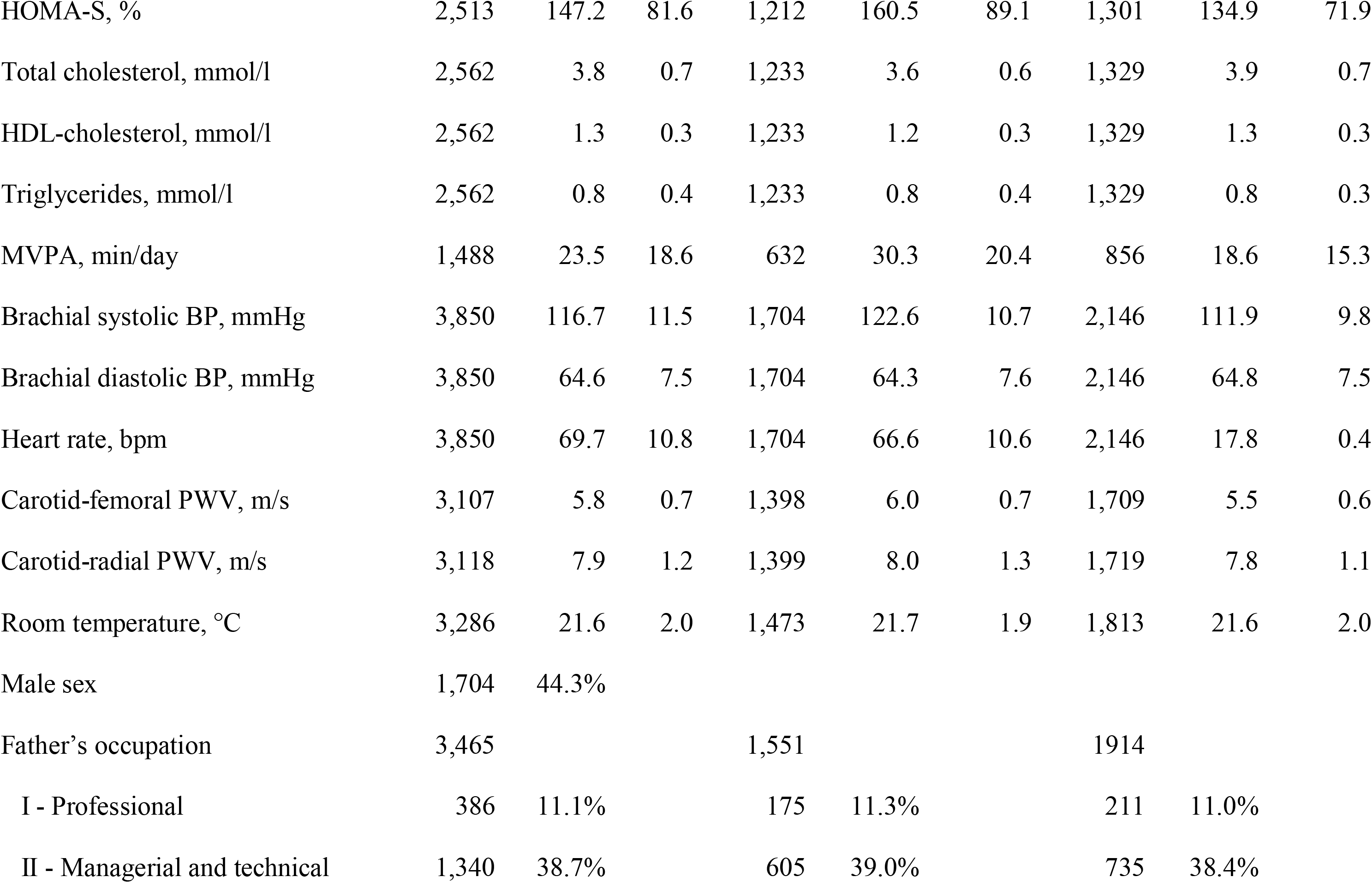
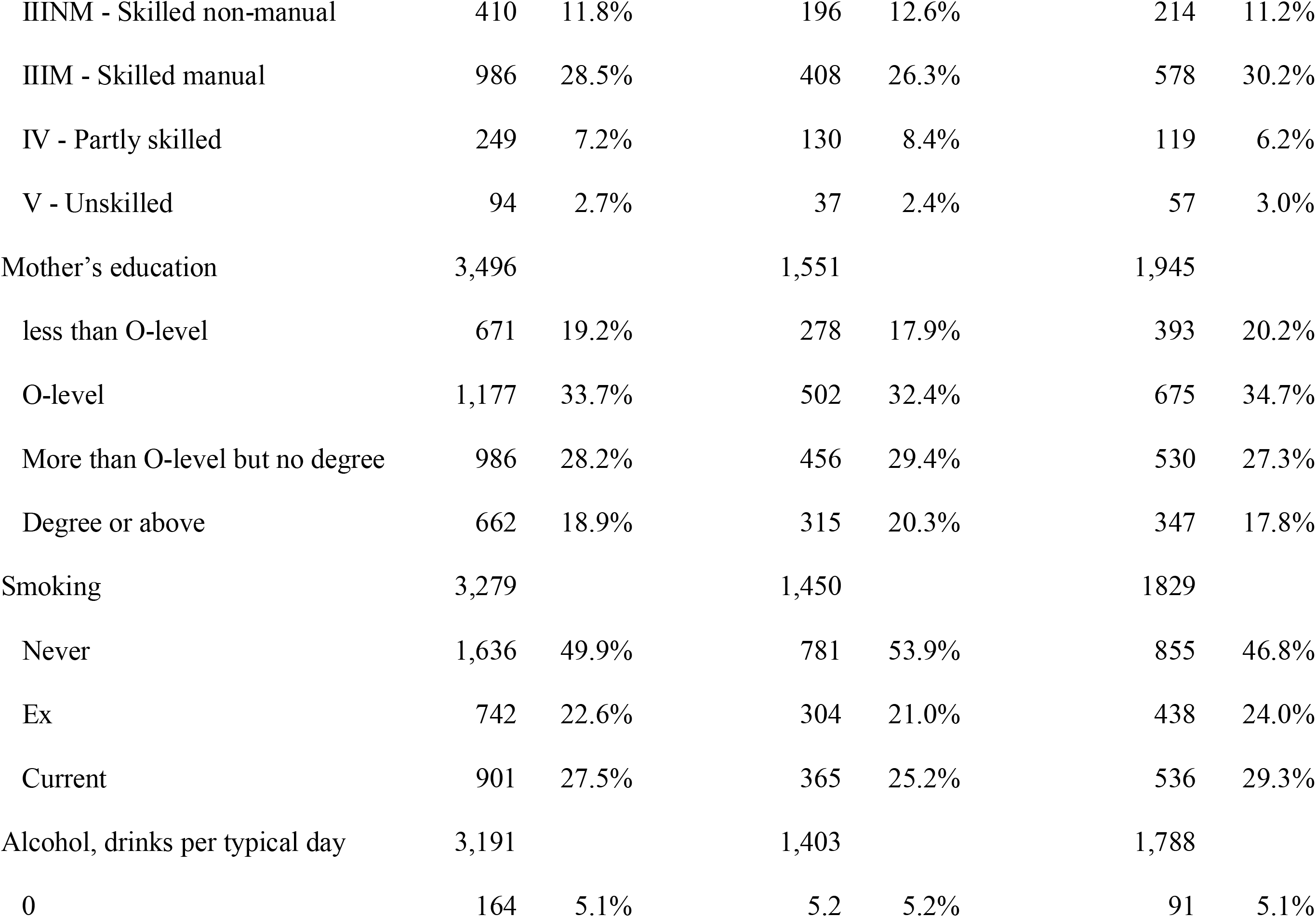
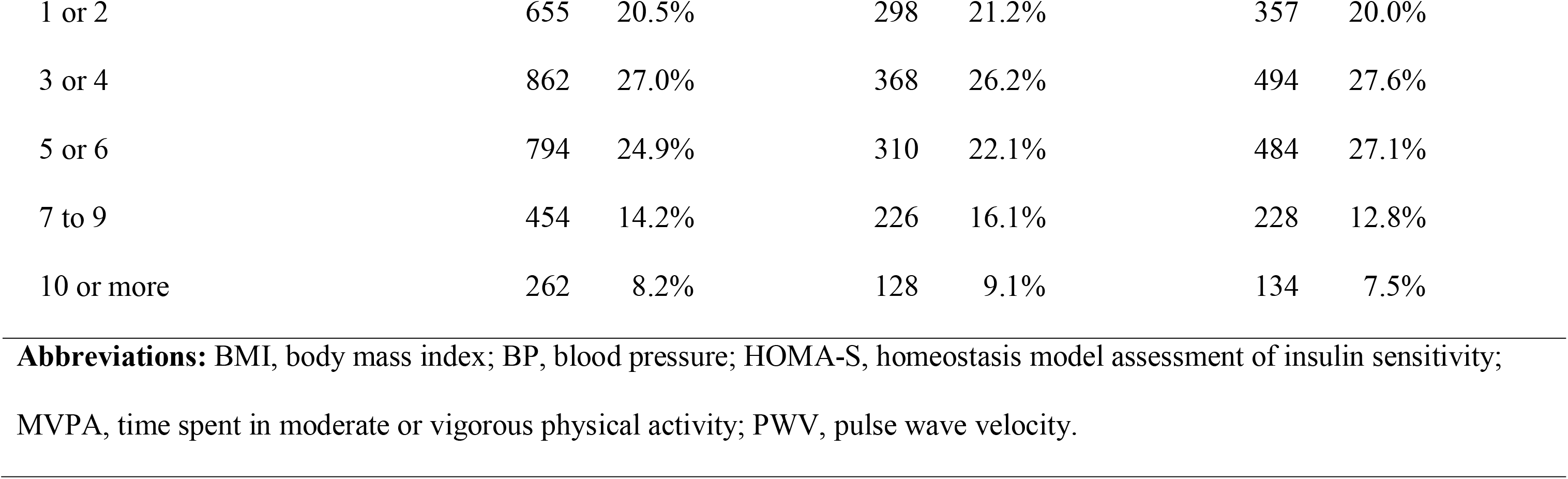
Participant characteristics.

| Variable | All |  |  | Male |  |  | Female |  |  |
| --- | --- | --- | --- | --- | --- | --- | --- | --- | --- |
|  | N | Mean/<br>% | SD | N | Mean/<br>% | SD | N | Mean/<br>% | SD |
| Age, y | 3,850 | 17.8 | 0.4 | 1,704 | 17.8 | 0.4 | 2,146 | 17.8 | 0.4 |
| Height, cm | 3,767 | 171.3 | 9.4 | 1,672 | 178.9 | 6.7 | 2,095 | 165.2 | 6.2 |
| Weight, kg | 3,771 | 67.0 | 13.5 | 1,675 | 72.3 | 12.9 | 2,096 | 62.9 | 12.4 |
| BMI, kg/m<sup>2</sup> | 3,767 | 22.8 | 4.0 | 1,672 | 22.6 | 3.7 | 2,095 | 23.0 | 4.2 |
| Fat mass, kg | 3,706 | 18.2 | 10.4 | 1,650 | 13.9 | 9.7 | 2,056 | 21.8 | 9.6 |
| Lean mass, kg | 3,706 | 45.7 | 10.0 | 1,650 | 55.3 | 6.1 | 2,056 | 38.0 | 4.3 |
| Fasting glucose, mmol/l | 2,562 | 5.0 | 0.4 | 1,233 | 5.1 | 0.41 | 1,329 | 4.9 | 0.4 |
| Insulin, pmol/l | 2,528 | 49.4 | 42.2 | 1,223 | 45.5 | 36.2 | 1,305 | 53.0 | 46.9 |
| HOMA-S, % | 2,513 | 147.2 | 81.6 | 1,212 | 160.5 | 89.1 | 1,301 | 134.9 | 71.9 |
| Total cholesterol, mmol/l | 2,562 | 3.8 | 0.7 | 1,233 | 3.6 | 0.6 | 1,329 | 3.9 | 0.7 |
| HDL-cholesterol, mmol/l | 2,562 | 1.3 | 0.3 | 1,233 | 1.2 | 0.3 | 1,329 | 1.3 | 0.3 |
| Triglycerides, mmol/l | 2,562 | 0.8 | 0.4 | 1,233 | 0.8 | 0.4 | 1,329 | 0.8 | 0.3 |
| MVPA, min/day | 1,488 | 23.5 | 18.6 | 632 | 30.3 | 20.4 | 856 | 18.6 | 15.3 |
| Brachial systolic BP, mmHg | 3,850 | 116.7 | 11.5 | 1,704 | 122.6 | 10.7 | 2,146 | 111.9 | 9.8 |
| Brachial diastolic BP, mmHg | 3,850 | 64.6 | 7.5 | 1,704 | 64.3 | 7.6 | 2,146 | 64.8 | 7.5 |
| Heart rate, bpm | 3,850 | 69.7 | 10.8 | 1,704 | 66.6 | 10.6 | 2,146 | 17.8 | 0.4 |
| Carotid-femoral PWV, m/s | 3,107 | 5.8 | 0.7 | 1,398 | 6.0 | 0.7 | 1,709 | 5.5 | 0.6 |
| Carotid-radial PWV, m/s | 3,118 | 7.9 | 1.2 | 1,399 | 8.0 | 1.3 | 1,719 | 7.8 | 1.1 |
| Room temperature, °C | 3,286 | 21.6 | 2.0 | 1,473 | 21.7 | 1.9 | 1,813 | 21.6 | 2.0 |
| Male sex | 1,704 | 44.3% |  |  |  |  |  |  |  |
| Father's occupation | 3,465 |  |  | 1,551 |  |  | 1914 |  |  |
| I - Professional | 386 | 11.1% |  | 175 | 11.3% |  | 211 | 11.0% |  |
| II - Managerial and technical | 1,340 | 38.7% |  | 605 | 39.0% |  | 735 | 38.4% |  |

|  |  |  |  |  |  |  |
| --- | --- | --- | --- | --- | --- | --- |
| IIINM - Skilled non-manual | 410 | 11.8% | 196 | 12.6% | 214 | 11.2% |
| IIIM - Skilled manual | 986 | 28.5% | 408 | 26.3% | 578 | 30.2% |
| IV - Partly skilled | 249 | 7.2% | 130 | 8.4% | 119 | 6.2% |
| V - Unskilled | 94 | 2.7% | 37 | 2.4% | 57 | 3.0% |
| Mother's education | 3,496 |  | 1,551 |  | 1,945 |  |
| less than O-level | 671 | 19.2% | 278 | 17.9% | 393 | 20.2% |
| O-level | 1,177 | 33.7% | 502 | 32.4% | 675 | 34.7% |
| More than O-level but no degree | 986 | 28.2% | 456 | 29.4% | 530 | 27.3% |
| Degree or above | 662 | 18.9% | 315 | 20.3% | 347 | 17.8% |
| Smoking | 3,279 |  | 1,450 |  | 1,829 |  |
| Never | 1,636 | 49.9% | 781 | 53.9% | 855 | 46.8% |
| Ex | 742 | 22.6% | 304 | 21.0% | 438 | 24.0% |
| Current | 901 | 27.5% | 365 | 25.2% | 536 | 29.3% |
| Alcohol, drinks per typical day | 3,191 |  | 1,403 |  | 1,788 |  |
| 0 | 164 | 5.1% | 5.2 | 5.2% | 91 | 5.1% |
| 1 or 2 | 655 | 20.5% | 298 | 21.2% | 357 | 20.0% |
| 3 or 4 | 862 | 27.0% | 368 | 26.2% | 494 | 27.6% |
| 5 or 6 | 794 | 24.9% | 310 | 22.1% | 484 | 27.1% |
| 7 to 9 | 454 | 14.2% | 226 | 16.1% | 228 | 12.8% |
| 10 or more | 262 | 8.2% | 128 | 9.1% | 134 | 7.5% |

**Abbreviations:** BMI, body mass index; BP, blood pressure; HOMA-S, homeostasis model assessment of insulin sensitivity; MVPA, time spent in moderate or vigorous physical activity; PWV, pulse wave velocity.

Primary analyses were performed using listwise deletion to handle missing data, on the assumption that conditional independence between missingness and aortic-to-brachial pulse pressure amplification (outcome) was more plausible than a missing-at-random assumption.

## Results

The participants’ characteristics are listed in Table 1. 66% of the participants were female, and the mean age for both sexes was 17.8 years. The mean brachial BP for both sexes was 116.7/64.6 mmHg, with males having a higher brachial BP than females. Compared with all those invited, participants who attended clinics were more likely to be female and come from more advantaged socioeconomic circumstances (Supplementary Table S1).

Figure 1A - D show a comparison of brachial and aortic systolic and diastolic BP in males and females. The distribution of differences in brachial and aortic systolic BP between males and females is shown in Supplementary Figure S2.

**Figure 1.**
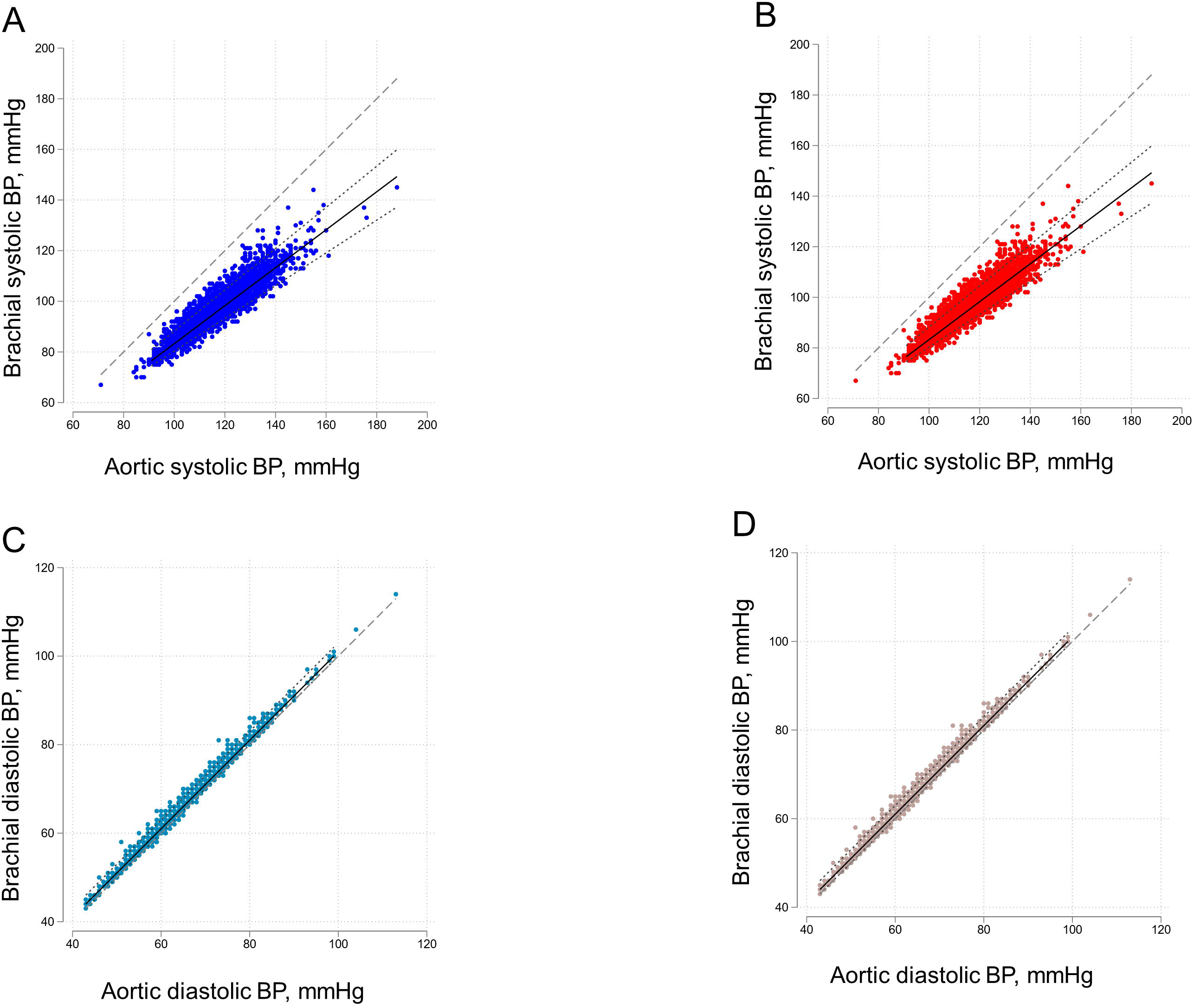
Relationship between brachial and aortic BP. A) Systolic BP (Male), B Systolic BP (Female), C) Diastolic BP (Male), D) Diastolic BP (Female). Lines are fractional polynomial fits (solid line) to the data with 5% and 95% quantile limits (dotted line). The dashed line is the line of unity.

For both males and females, there was a difference between aortic and brachial systolic BP (male: -22.3 [-31.2, -13.3] mmHg; female: -17.8 [-25.5, -10.0] mmHg). The difference between aortic and brachial systolic BP was attributable to large aortic to brachial amplification which was of similar magnitude in both sexes (male: median 1.71 (range 1.23 to 1.96); female: median 1.71 (range 1.10 to 2.08)), although because BP in males was higher absolute differences were larger for males than females. There was no evidence of a difference in the slope of the relationship between the aortic and brachial systolic BP (p = 0.13). Differences between SBP2, an alternative measure of aortic systolic BP and brachial systolic BP, were even larger (male: -33.7 [-53.0, -14.4] mmHg; female: -25.0 [-40.7, -9.2] mmHg), with larger differences between males and females.

In contrast, there was a minimal difference between aortic and brachial diastolic BP in both sexes (male: 1.4 [-0.4, 3.1] mmHg; female: 1.2 [-0.3, 2.8] mmHg) (Figure 1 C & D).

### Classification and prevalence of hypertension using brachial and aortic BP

On the basis of brachial BP, 123(3%) participants were classified as hypertensive; of these ISH accounted for 109(88%) were hypertensive. There was a marked sex difference in hypertension prevalence based on brachial BP, with 101 hypertensive males (6% of males) and 22 hypertensive females (1% of females). ISH accounted for 98(97%) cases of hypertension in males and only 11(50%) cases of hypertension in females.

When aortic BP was used, only 23 patients (0.6%) had aortic hypertension. Of these, five (21%) were due to aISH. A minimal sex difference was observed for aortic hypertension: 9 males (0.5%) had aortic hypertension, of which 4 were attributable to aISH. In contrast, 14 females (0.7%) had aortic hypertension, of which only one had aISH.

The classification matrices for aortic and brachial hypertension are presented in Table 2A. Although the sensitivity, specificity, and negative predictive value of brachial BP for aortic hypertension were excellent, brachial BP had a poor positive predictive value for aortic hypertension (13.8%).

**Table 2A.** Classification matrix for aortic and brachial SBP according to the ESH guideline classification.

| **Aortic BP** | **Brachial BP** |  | **Total** |
| --- | --- | --- | --- |
|  | Hypertensive | Normotensive |  |
| Aortic Hypertensive | 17 | 2 | 19 |
| Aortic Normotensive | 106 | 3725 | 3831 |
| Total | 123 | 3727 | 3850 |

| **Measure** | **Estimate** | **95% confidence interval** |
| --- | --- | --- |
| Sensitivity | 89.5% | (66.9%, 98.7%) |
| Specificity | 97.2% | (96.7%, 97.7%) |
| Likelihood ratio (+) | 32.3 | (25.0, 41.0) |
| Likelihood ratio (-) | 0.11 | (0.03, 0.40) |
| Positive predictive value | 13.8% | (8.3%, 21.2%) |
| Negative predictive value | 99.9% | (99.8%, 100%) |

**Table 2B.** Classification matrix for aortic and brachial ISH.

| **Aortic BP** | **Brachial BP** |  | **Total** |
| --- | --- | --- | --- |
|  | **ISH** | **No ISH** |  |
| Aortic ISH | 5 | 0 | 5 |
| Aortic No ISH | 104 | 3741 | 3845 |
| Total | 109 | 3741 | 3850 |

| **Measure** | **Estimate** | **95% confidence interval** |
| --- | --- | --- |
| Sensitivity | 100% | (47.8%, 100%) |
| Specificity | 97.3% | (96.7%, 97.8%) |
| Likelihood ratio (+) | 37.0 | (30.6, 44.7) |
| Likelihood ratio (-) | 0 | - - |
| Positive predictive value | 4.6% | (1.5%, 10.4%) |
| Negative predictive value | 100% | (99.9%, 100%) |

The results for ISH were similar (Table 2B) with excellent sensitivity, specificity, and negative predictive value of brachial BP for aortic ISH but very poor positive predictive value for aortic ISH (4.6%).

The results using a threshold of 130/80 mmHg for diagnosis of hypertension as recommended by the American Academy of Pediatrics Clinical Practice Guideline are shown in the Supplementary TableS2; these showed excellent sensitivity and negative predictive values but lower specificity (85.6%) and a poorer positive predictive value (4%) than the ESH-based criteria.

### Relationship of aortic hypertension compared with ISH on early target organ damage in adolescence

Both aortic and brachial hypertension without aortic hypertension were associated with cardiac target organ damage (higher LVMI), although the elevation in LVMI was larger in patients with aortic hypertension than in those with ISH (13.4 (95% CI, 4.0, 22.8) g/m^1.7^; p = 0.005 versus 7.1(95% CI, 1.4, 12.8) g/m^1.7^; p = 0.015) after adjustment for potential confounders. There was no convincing evidence that either brachial or aortic hypertension or ISH was associated with vascular target organ damage, as assessed by cIMT or cfPWV.

### Predictors of aortic to brachial pulse pressure amplification

For males 4 variables were identified as predictors of aortic to brachial pulse pressure amplification (age, heart rate, crPWV, and room temperature). The out-of-sample r^2^ was small (r^2^ = 0.04), indicating that it was largely unexplained by these variables. A sensitivity analysis excluding physical activity identified a total of eight predictors (age, total fat mass, HDL-cholesterol, heart rate, smoking, crPWV, cfPWV, and room temperature) that were only minimally different in terms of model fit (r^2^ = 0.08) despite the larger number of predictors and fewer missing values.

For females, five variables (HDL-cholesterol, triglycerides, heart rate, maternal education, and cfPWV) were predictors of amplification; however, the out-of-sample r^2^ was very modest (r^2^ = 0.03). Sensitivity analysis excluding physical activity only marginally improved the variance explained by the model (r^2^ = 0.08).

## Discussion

Brachial systolic BP was substantially higher than aortic systolic BP in both sexes in a birth cohort of adolescents and that this difference varied considerably between individuals. This difference in adolescents (male: 22mmHg, female: 18mmHg) was considerably higher than has been typically found in adults (∼12mmHg[13,37]). Higher brachial systolic BP was attributable to pulse pressure amplification and led to a large discrepancy in hypertension diagnosis when criteria based on brachial and aortic BP were compared.

Both aortic hypertension and brachial hypertension without aortic hypertension, which corresponded to ISH in almost all instances, were associated with increased LVMI, although the association was stronger with aortic hypertension and there was no evidence of vascular target organ damage (based on PWV or cIMT) in people with hypertension or ISH. Penalized regression (elastic net) using a wide range of measured variables, (including age, heart rate, cfPWV, crPWV, and CV risk factors) explained <10% of the variance in amplification in either sex, indicating that differences between brachial systolic BP and aortic systolic BP cannot be predicted, at least by these variables; therefore, amplification is an important and unexplained source of variation in brachial SBP. Our finding of large differences in brachial and aortic systolic BP due to amplification in young people is consistent with previous studies. In the Anglo-Cardiff Collaborative Trial II, younger age was by far the strongest predictor of amplification, with smaller contributions from sex, diabetes, smoking, and cardiovascular diseases.[13] Since the age range in our birth cohort was narrow and participants were younger and predominantly healthy our findings are broadly consistent with these observations.

Aortic to brachial pulse pressure amplification is attributable to wave reflection from the downstream circulation, which arises because of impedance mismatching in the peripheral arterial tree.[11] At an individual bifurcation, the magnitude of wave reflection depends on the geometrical and biophysical properties of the parent and daughter arteries,[38] but reflection patterns from anatomically asymmetric trees such as those in the upper limb are complex[39] which probably explains why the magnitude of pulse pressure amplification is unpredictable between individuals.

The high variability in amplification between individuals in this age group results in discrepancies in the identification of hypertension based on aortic or brachial BP. The definition of hypertension is inherently arbitrary,[40] and while it is relatively uncommon in children and adolescents (current global estimates are around 2-13%,[2,3]), its prevalence has risen sharply since the 1990’s and there are calls for more screening.[41] Previous studies have reported that aortic BP is a better predictor of outcomes than brachial BP in adults, and we have reported previously in ALSPAC that aortic pulse pressure is more closely associated with left ventricular hypertrophy than brachial pulse pressure in adolescence.[21] Another more recent study using ambulatory aortic BP in selected normotensive and hypertensive young people drew similar conclusions with regard to the superiority of aortic BP as a predictor of elevated LV mass.[42] While the evidence cannot be definitive in the absence of hard endpoints (which will take years to accrue), our data suggest that aortic BP may be a more appropriate indicator of risk in young people - this is consistent with most key organs being exposed to aortic, not brachial systolic BP. Since cuff-based devices are now available for measurement of aortic pressure, this suggests that their wider use in children and adolescents may support therapeutic decision-making.

Our findings of large variability in amplification may also help explain previous conflicting observations in young adults comparing the relationships of systolic and diastolic BP with later CV mortality,[7,43,44] since this variability is likely to introduce uncertainty in estimated risk relationships. Although aortic BP was a stronger predictor of early cardiac target organ damage than brachial BP, there was a weak association between ISH and higher LVMI in males. This is consistent with previous work showing associations between systolic hypertension and increased LVMI in children.[45]

Our study has several limitations. At recruitment, ALSPAC was not representative of the UK population,[22] and it may also differ from more contemporary cohorts. As with all cohort studies, there has been attrition over time, which may introduce further bias and limit the generalizability of the findings. Further studies in different geographic regions covering different age ranges during childhood would be valuable. In ALSPAC measurement of BP was made on only one occasion while diagnosis of hypertension usually requires BP measurements on multiple occasions. This may have led to a higher proportion of people identified as having BP in the hypertensive range. Nevertheless, it should not have influenced the difference between brachial and aortic pressure,[46] and the white coat effect is similar on brachial and aortic pressure, at least in adults.[47] The strengths of the study include its large sample size and the standardized measurements of BP and other cardiovascular indices.

## Conclusion

Blood pressure measured at the brachial artery overestimates aortic blood pressure in adolescents owing to marked aortic-to-brachial pulse pressure amplification. The use of brachial blood pressure could result in an overdiagnosis of hypertension during adolescence.

## Data Availability

Access to ALSPAC data is through a system of managed open access (http://www.bristol.ac.uk/alspac/researchers/access/).

## Acknowledgements

We are extremely grateful to all the families who took part in the study, the midwives for their help in recruiting them, and the ALSPAC team, which included interviews, computer and laboratory technicians, clerical workers, research scientists, volunteers, managers, receptionists, and nurses.

