## Supplementary data for "Differences Between Brachial And Aortic Blood Pressure In Adolescence and their implications for diagnosis of hypertension"

**Supplementary tables**

**Table S1.** Comparison of those invited, who did or did not participate in the aortic blood pressure study.

|  | Participated in aortic blood pressure study | | |
| --- | --- | --- | --- |
|  | No | Yes | P |
| N | 6,193 (61.4%) | 3,894 (38.6%) |  |
| Sex |  |  |  |
| Male | 3,267 (52.8%) | 1,723 (44.2%) | <0.001 |
| Female | 2,926 (47.2%) | 2,171 (55.8%) |  |
| Social class at 17y |  |  |  |
| I - Professional | 395 (7.6%) | 388 (11.1%) | <0.001 |
| II - Managerial and   technical | 1,715 (32.8%) | 1,351 (38.6%) |  |
| IIINM - Skilled non-  manual | 545 (10.4%) | 413 (11.8%) |  |
| IIIM - Skilled manual | 1,997 (38.2%) | 999 (28.6%) |  |
| IV - Partly skilled | 412 (7.9%) | 253 (7.2%) |  |
| V - Unskilled | 167 (3.2%) | 95 (2.7%) |  |
| Mother's highest educational qualification |  |  | <0.001 |
| less than O-level | 1,616 (30.5%) | 679 (19.2%) |  |
| O-level | 1,863 (35.2%) | 1,193 (33.8%) |  |
| A-level | 1,182 (22.3%) | 996 (28.2%) |  |
| degree or above | 633 (12.0%) | 665 (18.8%) |  |

**Table S2** **Classification matrix for aortic and brachial SBP according to the AAPCP guideline classification**

|  | **Brachial BP** | |  |
| --- | --- | --- | --- |
| **Aortic BP** | Hypertensive | Normotensive | Total |
| Aortic Hypertensive | 23 | 0 | 23 |
| Aortic Normotensive | 551 | 3276 | 3827 |
| Total | 574 | 3276 | 3850 |

| **Measure** | **Estimate** | **95% confidence interval** | |
| --- | --- | --- | --- |
| Sensitivity | 100% | (85.2%, | 100%) |
| Specificity | 85.6% | (84.4%, | 86.7%) |
| Likelihood ratio (+) | 7.0 | (6.4, | 7.5) |
| Likelihood ratio (-) | 0 | - | - |
| Positive predictive value | 4.0% | (2.6%, | 6.0%) |
| Negative predictive value | 100% | (99.9%, | 100%) |

**Supplementary figures**

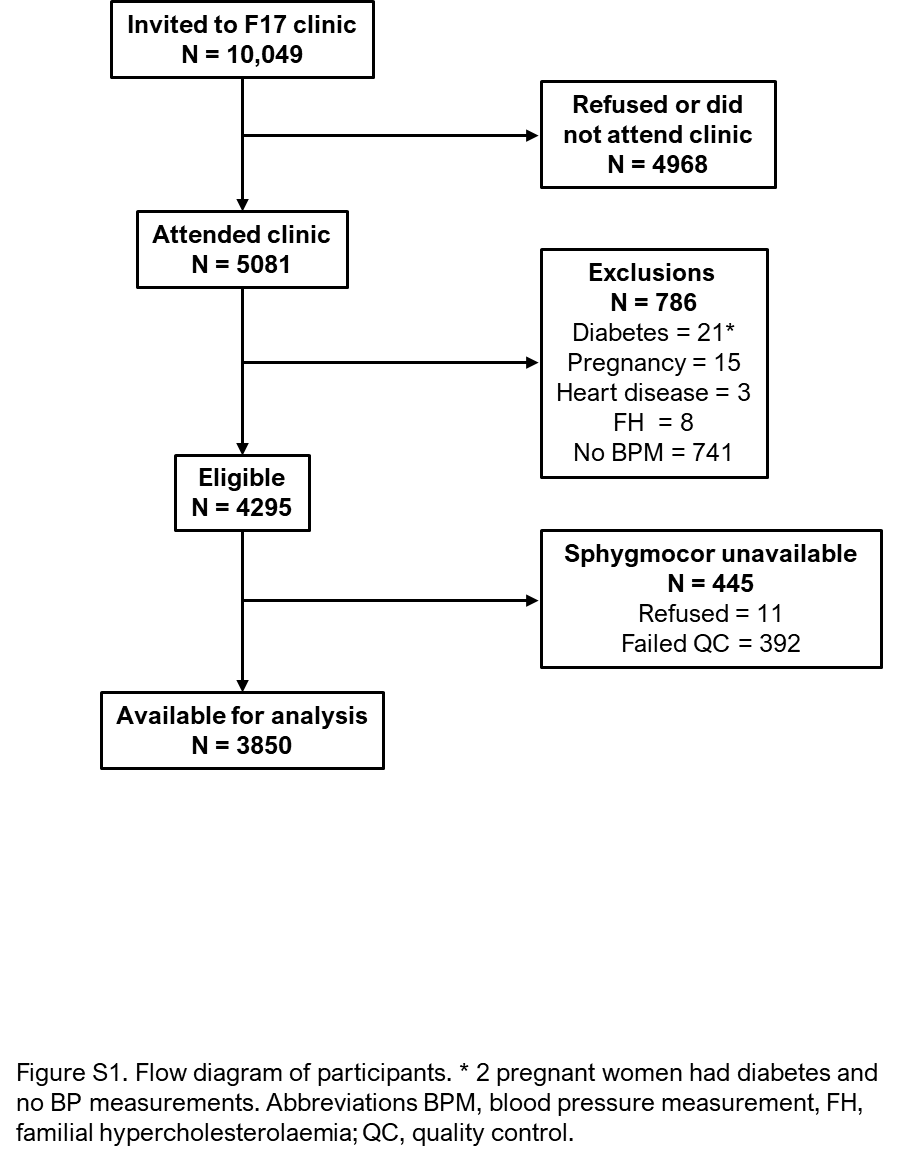

**Figure S1**. Flow diagram of participants. * 2 pregnant women had diabetes and no blood pressure measurements. Abbreviations BPM, blood pressure measurement, FH, familial hypercholesterolaemia; QC, quality control.

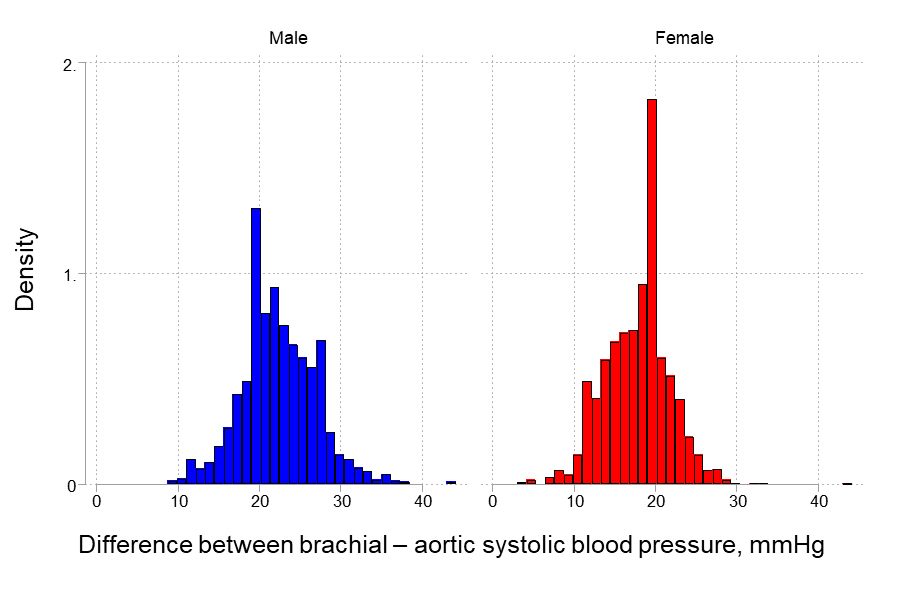

**Figure S2:** The difference between brachial-aortic systolic blood pressure in males and females.
